# Identifying High-Risk Adolescents for Mental Health Difficulties: A Machine Learning Analysis of the Health Behaviour in School-aged Children Study Across 46 Countries

**DOI:** 10.1101/2025.09.10.25335503

**Authors:** Shanquan Chen, Chengxu Long, Yuanxuan Cai, Sixian Du, Yiqiong Yang, Yunfei Li, Jianping Lu, Gordon Liu, Quinette Abegail Louw, Michael Ni

**Author notes:** **Corresponding author: Shanquan Chen, assistant professor, PhD**, International Centre for Evidence in Disability, London School of Hygiene & Tropical Medicine, London, United Kingdom, WC1E 7HT, School of Public Health, Li Ka Shing Faculty of Medicine, The University of Hong Kong.

## Abstract

**Background:** Adolescent mental health represents a global public health crisis, yet traditional surveillance methods lack the scalability and predictive power needed for effective early identification. This study aimed to develop and validate a machine learning framework to predict multiple psychosomatic health complaints (MPHC), a key indicator of mental distress, using large-scale, multi-national population survey data.

**Methods:** Data were drawn from the 2017–2018 Health Behaviour in School-aged Children (HBSC) survey, comprising an analytical sample of 225,421 adolescents aged 11, 13, and 15 years from 46 countries. The primary outcome was daily MPHC. The dataset was partitioned for training (80%) and testing (20%), with eight distinct machine learning algorithms developed and evaluated. Analyses were stratified by sex and school grade. Shapley Additive Explanations (SHAP) analysis was applied to the optimal models to identify the most important predictors of mental health risk.

**Results:** Machine learning models demonstrated robust discriminatory performance for identifying adolescents with daily MPHC, with area under the receiver operating characteristic curve (ROC) values ranging from 0.76 to 0.79 across subgroups. Model performance peaked for girls in Grade 7 (AUC = 0.79; 95% CI: 0.78–0.80). SHAP analysis revealed that modifiable psychosocial factors were the most powerful predictors. High academic pressure, problematic social media use, and low family support consistently emerged as the top predictors across all subgroups. The analysis uncovered distinct, sex-differentiated risk architectures: for boys, physical fighting was a uniquely persistent and high-impact predictor across all grades, while for girls, difficulties with parental communication and academic pressure were particularly salient, especially during mid-adolescence.

**Conclusions:** Machine learning applied to standardized population survey data offers a scalable, accurate paradigm for predictive public health screening of adolescents at mental health risk. The findings challenge a “one-size-fits-all” approach, providing a data-driven mandate for the design of developmentally-timed and sex-specific interventions that target the distinct psychosocial risk factors shaping adolescent well-being.

## Introduction

Adolescent mental health has emerged as a paramount global public health challenge, with current epidemiological evidence indicating that one in seven adolescents (14%) aged 10-19 years experience mental health conditions worldwide[1]. The scope of this crisis has intensified considerably, with depression prevalence among adolescents increasing from 12.9% pre-pandemic to 25.2% and anxiety symptoms rising from 11.6% to 20.5%[2]. These increases demonstrate clear demographic disparities, with depression prevalence in adolescent females (26.5%) more than double that of males (12.2%), and pronounced socioeconomic gradients where depression prevalence decreases from 22.1% in families below poverty level to 7.4% in high-income families[3]. The developmental period of adolescence represents a critical neurobiological window characterised by heightened vulnerability to environmental stressors that collectively contribute to psychological distress.

Multiple psychosomatic health complaints (MPHC), encompassing the co-occurrence of psychological symptoms such as irritability and mood disturbances alongside somatic manifestations including headaches and sleep difficulties, have gained recognition as important early indicators of adolescent mental health burden. Psychosomatic symptoms occur in approximately 48% of children and adolescents, with Irish school-based studies documenting that irritability/bad temper (43.0%) and headache (26.0%) were the most commonly reported psychological and somatic symptoms among adolescents[4, 5]. Longitudinal evidence demonstrates robust predictive validity, with Swedish cohort studies showing that psychosomatic complaints in adolescence were clearly and consistently associated with increased likelihood of subsequent depression and anxiety symptoms in young adulthood[6].

The contemporary digital landscape has fundamentally transformed adolescent social ecology, introducing novel risk factors that significantly influence mental health trajectories. Research demonstrates consistent associations between social media use and increased risk of depression, anxiety, and internalizing problems among adolescents, with longitudinal studies confirming that social media use predicts higher depressive symptoms and greater internalizing problems over time[7, 8]. However, the association demonstrates complexity, as extremely low screen time also correlates with worse mental health, suggesting optimal moderate use patterns[9]. The ubiquity of digital media among adolescents necessitates comprehensive assessment frameworks that can simultaneously evaluate traditional psychosocial determinants alongside contemporary digital environmental factors.

Traditional approaches to adolescent mental health surveillance have relied predominantly on clinical assessment protocols within healthcare settings, facing substantial limitations including delayed help-seeking behaviours and limited service accessibility. For instance, in the United States (US), no comprehensive surveillance system exists for children’s mental health despite mental disorders affecting approximately one in five children and adolescents annually[10]. Up date to 2020, at most 20% of schools in the US engaged in mental health screening, highlighting significant gaps in early identification capabilities that could prevent progression to severe psychiatric disorders[11]. Systematic reviews identify 210 different mental health indicators across OECD countries, highlighting the fragmented surveillance landscape where 27.7% of effective indicators require integrating multiple data sources[12].

Machine learning methodologies offer transformative potential for advancing adolescent mental health research through their capacity to identify complex, non-linear relationships among multiple predictor variables[13, 14]. Recent applications have demonstrated clinically relevant performance, with depression and anxiety prediction models achieving area under the curve values of 0.78-0.81 using clinical data[15], while suicide risk prediction models demonstrate even higher performance with ranges of 0.81-0.86[16]. Digital biomarkers from smartphones show exceptional promise, with meta-analyses revealing pooled performance metrics reaching 89% accuracy, 87% sensitivity, and 93% specificity for depression detection[13].

However, critical knowledge gaps persist in the current literature. Many studies remain underpowered, with sample sizes limiting generalisability to broader adolescent populations[13]. Furthermore, few investigations have systematically examined the precision of machine learning approaches for predicting adolescent mental health outcomes using large-scale population-based data, or identified the most important predictor variables across comprehensive risk factor domains.

This study aims to address these critical knowledge gaps by applying a comprehensive machine learning framework to predict multiple psychosomatic health complaints among adolescents using data from the 2017-2018 Health Behaviour in School-aged Children (HBSC) survey across 46 countries. Leveraging this unprecedented dataset of over 225,000 adolescents, we employed eight distinct machine learning algorithms with stratification by biological sex and school grade to ensure findings appropriately reflect population heterogeneity. Specifically, this research seeks to answer three primary questions: first, what is the discriminatory performance and clinical utility of machine learning algorithms for predicting daily multiple psychosomatic health complaints among adolescents across diverse demographic subgroups; second, what are the most important modifiable predictor variables for identifying adolescents at highest risk for severe mental health difficulties, as determined through Shapley Additive Explanations (SHAP) analysis; and third, how do sex-differentiated and developmentally-sensitive risk architectures emerge across sociodemographic, psychosocial, environmental, and digital media domains throughout early-to-late adolescence. By addressing these questions, this investigation aims to advance both theoretical understanding of adolescent mental health determinants and practical applications for population-level screening and targeted intervention strategies.

## Methods

### Data source and participants

This investigation utilised data from the Health Behaviour in School-aged Children (HBSC) study, a World Health Organization-affiliated multinational research initiative that systematically assesses health, wellbeing, and behavioural patterns among adolescents across Europe and North America. The HBSC employs a sophisticated cross-sectional design, collecting data quadrennially from nationally representative samples of students aged 11, 13, and 15 years across participating countries. The survey methodology incorporates a complex probability sampling framework utilising stratification and primary sampling units to ensure robust population representativeness, with stratification parameters including age cohorts, geographical regions, and educational institutions to achieve coverage of at least 95% of the eligible target population within each sampling frame[17].

Primary sampling units, typically comprising schools or individual classrooms, are selected through systematic or random sampling procedures from comprehensive sampling frames of eligible educational institutions[17]. The HBSC protocol incorporates multiple methodological safeguards to minimise response bias, including strict anonymity protocols, standardised back-translated questionnaires, and supervised classroom administration procedures[17]. The survey instrument undergoes rigorous validation through pilot testing and incorporates post-collection data quality assessments to identify and mitigate potential sources of bias.

The present analysis examined data from the most recent publicly available HBSC survey cycle conducted during 2017-2018, as this wave contained extensive information on digital media use patterns that are central to contemporary adolescent mental health research[18]. Response rates consistently exceeded 60% across participating countries, with detailed methodological specifications documented in previous publications[17]. The utilisation of secondary de-identified data exempted this investigation from institutional review board oversight, while all original HBSC surveys received appropriate ethical approval and informed consent from participants and guardians within respective jurisdictions.

### Outcome measurement

The primary outcome, multiple psychosomatic health complaints (MPHC), was assessed using a validated eight-item psychosomatic symptom inventory measuring the frequency of psychological and somatic manifestations including depressed mood, irritability, nervousness, sleep disturbances, dizziness, headache, abdominal pain, and back pain experienced during the preceding six months. Participants indicated symptom frequency on a five-point ordinal scale ranging from daily occurrence to rare or absent presentation. This measurement instrument has demonstrated robust psychometric properties through qualitative validation interviews with adolescent participants and quantitative test-retest reliability assessments yielding satisfactory intraclass correlation coefficients ranging from 0.61 to 0.75[19].

Multiple psychosomatic health complaints was operationally defined as the co-occurrence of two or more psychosomatic symptoms experienced at frequencies of “more than once weekly” or greater(4). This definitional approach was selected over sum-score methodologies for several methodological considerations. The frequency-based threshold provides superior conceptual alignment with clinically meaningful presentations of psychological distress by operationalising mental health burden as persistent, co-occurring symptomatology rather than cumulative symptom counts that may conflate frequent severe symptoms with numerous mild complaints. Furthermore, this definition ensures methodological consistency with established HBSC international research protocols, facilitating direct comparison with published literature and World Health Organization surveillance reports[20].

In the present study population, the prevalence of multiple psychosomatic health complaints occurring at least weekly was approximately 35%, while daily occurrence affected approximately 15% of participants. The latter prevalence closely approximates established rates of depression and anxiety disorders in paediatric populations. Given this study’s objective to identify adolescents at highest risk for severe mental health difficulties, we employed the more stringent criterion of daily symptom occurrence to enhance specificity for clinically significant presentations requiring targeted intervention.

### Potential predictors

The selection of potential predictors was guided by the ecological systems theory of human development and the biopsychosocial model of adolescent mental health. These complementary frameworks recognise that adolescent psychological wellbeing emerges from complex interactions between individual characteristics, interpersonal relationships, and broader environmental contexts. The ecological perspective emphasises how multiple nested systems— from microsystems such as family and peer relationships to macrosystems including socioeconomic structures—collectively influence developmental outcomes. The biopsychosocial model further delineates how biological factors, psychological processes, and social determinants converge to shape mental health trajectories during the critical developmental period of adolescence.

### Sociodemographic and Socioeconomic Determinants

Sociodemographic characteristics included chronological age, biological sex, and family composition variables encompassing parental cohabitation status and sibling presence. Socioeconomic position was assessed through the Family Affluence Scale III (FAS-III), a validated proxy measure of household socioeconomic status yielding scores from 0 to 13, where higher values indicate greater material affluence[21-23]. Perceived academic pressure was measured using established HBSC items.

### Anthropometric and Health Behaviour Variables

Body mass index was calculated from self-reported height and weight measurements and standardised according to World Health Organization age- and sex-specific reference standards to generate z-scores (zBMI)[24]. Participants were subsequently categorised into weight status groups including thinness (zBMI ≤ -2), underweight (-1.99 to -1), normal weight (-0.99 to 0.99), overweight (1 to 1.99), and obesity (zBMI ≥ 2)[25]. Body perception was assessed through self-rated body shape evaluation on a five-point scale ranging from “much too thin” to “much too fat.”

Physical activity engagement was quantified through two complementary measures: weekly frequency of moderate-to-vigorous physical activity achieving at least 60 minutes duration, and leisure-time exercise frequency sufficient to induce perspiration or breathlessness[26, 27]. Substance use behaviours were assessed through standardised frequency scales measuring tobacco smoking, alcohol consumption, and cannabis use during the preceding 30-day period, with response categories ranging from never to daily use.

### Psychosocial and Environmental Factors

Social support was measured using validated subscales from the Multidimensional Scale of Perceived Social Support, specifically examining family support (exemplified by items such as “Family members really try their best to help me”) and peer support (including items such as “My friends really try to help me”)[28, 29]. Each domain comprised four items rated on seven-point Likert scales from “totally disagree” to “totally agree,” with mean scores calculated for each domain demonstrating excellent internal consistency (Cronbach’s α = 0.93 for both domains).

Communication patterns were evaluated through assessments of perceived accessibility of family members for discussing personal concerns, with separate evaluations for biological and step-parents using four-point scales ranging from “very easy” to “very difficult.” Digital communication preferences were measured using three items from the Perceived Depth of Online Communication Scale, assessing participants’ perceptions of online versus offline communication effectiveness for intimate disclosure[29].

Social media engagement was characterised through two distinct constructs: intensive social media use, defined as maintaining online contact “almost all the time throughout the day” with at least one category of social contacts[30, 31], and problematic social media use, measured using the nine-item Social Media Disorder Scale (α = 0.89))[32]. Problematic use was defined as endorsement of six or more addiction-like symptoms including preoccupation, withdrawal, tolerance, and functional impairment[30].

School environmental factors encompassed subjective school satisfaction, perceived academic pressure, and school climate assessments[33]. Teacher-related climate was evaluated through three items measuring acceptance, care, and trust perceptions (α = 0.87), while student climate was assessed through three items evaluating peer kindness, acceptance, and social cohesion (α = 0.81) [33]. Both scales utilised five-point Likert response formats.

Additional risk factors included exposure to interpersonal violence through physical altercations, injury requiring medical attention, victimisation through traditional and cyber-bullying[34, 35], and adverse family circumstances including foster care placement.

### Statistics and Modeling

Cases exhibiting substantial missing data, defined as absence of information for 30% or more variables of interest, were excluded from analytical procedures. For remaining cases, multiple imputation using chained equations was implemented to generate imputed datasets, incorporating all variables of interest as predictors within the imputation model to preserve statistical relationships and maintain analytical power.

The complete dataset was randomly partitioned into training (80%) and testing (20%) subsets using stratified sampling to ensure balanced outcome distribution across both partitions. To address class imbalance within the training dataset, synthetic minority oversampling technique (SMOTE) was applied to generate synthetic samples for the minority class, thereby improving model performance and reducing bias towards majority class predictions[36].

Feature selection was conducted using recursive feature elimination with cross-validation (RFE-CV) implemented through the caret package’s caretFuncs algorithm[37]. This model-agnostic approach systematically evaluated predictor importance through iterative variable removal, with optimal feature sets determined through 5-fold cross-validation within the training dataset and final selection based on maximum receiver operating characteristic (ROC) values in the independent test dataset.

Eight distinct machine learning algorithms were implemented to ensure methodological robustness and identify optimal predictive approaches across different algorithmic families. Linear approaches included linear discriminant analysis (using the “sda” method) and logistic regression (using “glm” with the binomial family), while non-linear methods encompassed extreme gradient boosting trees (using “xgbTree”), k-nearest neighbours (using “knn”), neural networks (using “nnet”), and naive Bayes classifiers (using “naive_bayes”). Advanced ensemble methods comprised support vector machines with radial basis functions (using “svmRadial”) and random forest algorithms (using “rf”). All models were implemented using the ‘caret’ R package and underwent hyperparameter optimisation through 5-fold cross-validation during training phases.

Model performance was evaluated exclusively on the reserved test dataset using ROC metric, supplemented by comprehensive performance indicators including sensitivity, specificity, accuracy, area under the curve (AUC), F-score, Cohen’s kappa statistic, precision, and recall measures. To enhance model interpretability and clinical utility, Shapley Additive Explanations (SHAP) analysis was applied to the optimal-performing model, providing transparent visualisation of individual feature contributions to prediction outcomes.

Recognising established gender and developmental differences in adolescent mental health presentations, all analytical procedures were stratified by biological sex and school grade to ensure findings appropriately reflect population heterogeneity.

A sensitivity analysis was conducted with “at least weekly” as the cut-off to assess the robustness of our findings using a less stringent threshold for multiple psychosomatic health complaints (MPHC).

Statistical analyses were conducted using R version 4.3 with appropriate packages for machine learning implementation and model evaluation.

## Results

### Characteristics of the Study Population

The analytical sample comprised 225,421 adolescents from 46 countries, with 15.5% (n = 34,834) experiencing daily multiple psychosomatic health complaints (MPHC). Comprehensive descriptive analysis revealed substantial disparities between groups (**Table 1**). Girls were significantly overrepresented in the MPHC group (64.5% vs 48.8%; SMD = -0.32), with affected adolescents being slightly older (13.75 vs 13.48 years; SMD = -0.17) and reporting lower family affluence (7.51 vs 7.99; SMD = 0.17).

The most pronounced differences emerged within psychosocial domains. Adolescents with MPHC reported markedly reduced social support from family (SMD = 0.44), peers (SMD = 0.43), and teachers (SMD = 0.40), alongside substantially greater parental communication difficulties (father SMD = 0.48; mother SMD = 0.44). Exposure to adverse interpersonal experiences was significantly elevated, with frequent traditional bullying affecting 9.2% versus 2.6% of controls, and similar disparities observed for cyberbullying, physical fighting, and medically-attended injuries.

Academic and digital environmental factors demonstrated strong imbalances. High academic pressure affected 29.5% of the MPHC group compared with 10.3% of controls (SMD = 0.56), while problematic social media use scores were substantially elevated (2.67 vs 1.61; SMD = - 0.47). Among MPHC group, health behaviours revealed higher substance use rates including daily smoking (4.2% vs 1.3%) and frequent alcohol consumption. Physical activity patterns were mixed among MPHC group, with lower moderate-to-vigorous activity but higher out-of-school exercise frequency.

**Sup table 1** compared baseline characteristics between complete and missing data cases to assess selection bias. Some variables showed imbalance, but standardized mean differences remained within acceptable analytical thresholds.

### Discriminatory Performance of Machine Learning Models

Figure 1. presents ROC curves for optimal machine learning models predicting multiple psychosomatic health complaints across demographic subgroups. Models demonstrated robust predictive performance with ROC values ranging from 0.76-0.79. Among girls, performance varied slightly across grades: 0.77 (95% CI: 0.76-0.78) in Grade 5, 0.79 (95% CI: 0.78-0.80) in Grade 7, and 0.77 (95% CI: 0.76-0.78) in Grade 9, with Grade 7 achieving peak performance.

Boys showed consistent performance: 0.77 (95% CI: 0.76-0.78) in Grade 5, 0.76 (95% CI: 0.75-0.77) in Grade 7, and 0.76 (95% CI: 0.75-0.78) in Grade 9. Other performance indicators can be found in **Sup Table 2**.

### The Architecture of Adolescent Mental Health Risk

To enhance interpretability and generate clinically actionable insights, Shapley Additive Explanations (SHAP) analysis was applied to optimal models. This technique quantifies individual feature contributions to risk predictions. **Figure 2** visualises SHAP analysis where point position indicates risk impact (positive increases risk, negative decreases) and colour represents feature magnitude (red=high, blue=low). **Figure 3** ranks the top 15 predictors by importance for each demographic subgroup, with red bars denoting risk factors and blue bars representing protective factors. Analysis revealed consistent high-impact predictors across subgroups, primarily academic pressure, problematic social media use, and family support quality. However, hierarchical structure and developmental trajectories demonstrated significant sex-specific differences.

### Sex-Differentiated Risk Profiles

SHAP analysis revealed distinct risk architectures reflecting known sex differences in adolescent psychopathology (**Figure 3**). Girls demonstrated relatively stable risk profiles across adolescence. Academic pressure dominated across Grades 5, 7, and 9 (importance +0.056 to +0.063), followed consistently by problematic social media use (+0.047 to +0.051). Family support and affluence emerged as primary protective factors. Conversely, boys exhibited dynamic predictor hierarchies. In Grade 5, problematic social media use was most important (+0.049), but its influence diminished with age while academic pressure became dominant by Grade 9 (+0.062). A crucial distinction from the female profile was physical fighting’s persistent high importance across all male grades (+0.034 to +0.036).

### Developmental Shifts in Risk and Protective Factors

Predictor trajectory analysis (**Figure 3**) revealed complex, evolving risk architectures throughout adolescence. Familial communication patterns demonstrated particular dynamism. Among girls, difficulty talking to mothers nearly doubled between Grades 5-7 (+0.012 to +0.023) before receding in Grade 9, identifying mid-adolescence as a critical vulnerability period. Boys showed similar but less pronounced father-communication patterns. Contrasting this volatility, family support maintained stable protective importance for both girls (+0.023 to +0.026) and boys (+0.019 to +0.031) across all grades. Behavioural risk factors displayed clear developmental patterns. Substance use importance increased markedly in older adolescents of both sexes, reflecting greater prevalence and distress signalling potential. Physical fighting maintained consistently high importance for boys (+0.034 to +0.036) but remained less prominent for girls across all developmental stages.

The sensitivity analysis with “at least weekly” as the cut-off (**Sup Table 3** and **Sup Figure 1 to 3**) confirmed the primary findings, including model performance, top predictive factors, and developmental shifts.

## Discussion

### Principal findings

In this analysis of 225,421 adolescents from 46 countries participating in the Health Behaviour in School-aged Children study, we demonstrated that machine learning approaches can effectively identify adolescents at highest risk for multiple psychosomatic health complaints (MPHC) with robust discriminatory performance across demographic subgroups. Our models achieved ROC values ranging from 0.76 to 0.79, with girls demonstrating peak performance in Grade 7 (ROC = 0.79) and boys maintaining consistent performance across developmental stages. SHAP analysis revealed that psychosocial and environmental factors substantially outperformed demographic or anthropometric indicators in risk prediction, with academic pressure emerging as the dominant predictor across older cohorts of both sexes (importance values +0.056 to +0.063). Problematic social media use maintained consistently high predictive importance, particularly among girls (+0.047 to +0.051), while demonstrating declining influence among boys from Grade 5 to Grade 9. Family support emerged as the most stable protective factor across all demographic subgroups (+0.019 to +0.031), contrasting with the dynamic trajectories observed for peer-related and behavioural variables. Sex-differentiated risk profiles were evident, with physical fighting representing a persistent high-impact predictor uniquely among boys (+0.034 to +0.036), while communication difficulties with parents showed pronounced importance among girls, particularly during mid-adolescence. These findings underscore the potential utility of machine learning approaches for population-level screening while highlighting the necessity for developmentally-tailored and sex-specific intervention strategies targeting modifiable psychosocial risk factors.

### Explanation and compare with previous studies

The present study achieved robust discriminatory performance (ROC 0.76-0.79) across 225,000 adolescents from 46 countries, establishing a new benchmark for scalable mental health screening. This performance matches or exceeds previous population-based models, including a Swedish twin study (ROC 0.739)[38], and approaches that of resource-intensive clinical data sources like electronic health records (AUC 0.78-0.81)[15]. While neuroimaging-based models achieve superior performance (AUC >0.90)[39, 40], their prohibitive costs preclude widespread implementation. The key contribution lies not in the specific ROC values, but in demonstrating that standardized survey instruments can achieve comparable accuracy across diverse international settings. This addresses critical limitations of prior research confined to single populations or clinical contexts[38], validating machine learning’s feasibility for global surveillance using population-level data. The peak performance in Grade 7 girls (ROC 0.79) identifies a critical developmental vulnerability window. This period (ages 11-13) coincides with pubertal onset, the emerging gender gap in depression, and heightened social media sensitivity[7, 8, 41-44], suggesting risk factors become more systematically patterned and predictable in this subgroup. These findings provide data-driven rationale for targeting early screening and interventions toward girls during this crucial developmental transition.

A central finding of this investigation is the decisive primacy of modifiable psychosocial and behavioral factors, as revealed SHAP analysis, over fixed demographic or anthropometric indicators in predicting adolescent mental distress, aligning with research emphasizing environmental and internal resources’ critical role in mental health trajectories[45]. Academic pressure emerged as the dominant predictor in older cohorts (SHAP importance +0.056 to +0.063), consistent with studies identifying it as a major contemporary stressor[46]. Problematic social media use showed consistently high importance for girls (+0.047 to +0.051), corroborating research linking behavioral addiction patterns to negative mental health outcomes[8, 47, 48]. Family support was the most stable protective factor across subgroups (+0.019 to +0.031), reinforcing meta-analytic evidence positioning secure familial relationships as central to adolescent resilience[45, 49, 50]. The novel contribution of this study lies in large-scale, quantitative hierarchization of risk factors across 46 countries. SHAP analysis provides data-driven ranking of relative impact, moving beyond risk factor cataloging to evidence-based prioritization for public health policy. This hierarchy suggests interventions targeting academic stress, digital well-being, and family cohesion would yield optimal population-level benefits. Cross-cultural consistency of top predictors indicates a “globalized signature” of adolescent stress driven by convergent pressures from standardized educational systems and ubiquitous digital platforms[48], suggesting core intervention targets may be similar across high- and middle-income nations.

This study reveals mental health risk architecture as dynamic, profoundly shaped by biological sex and developmental stage, uncovering distinct etiological pathways. These sex-differentiated and developmentally dynamic architectures align with literature documenting gender differences in adolescent mental health trajectories, including analysis of 566,829 adolescents across 73 countries confirming ubiquitous cross-cultural patterns: girls showing higher internalizing disorders, boys more externalizing problems[51]. The female risk profile—characterized by stable, high importance of academic pressure and problematic social media use—represents the internalizing pathway (anxiety, depression, social comparison) prevalent in females[41-43, 52]. Pronounced parent-child communication difficulties during mid-adolescence further support this, echoing research highlighting relational quality’s unique salience for female mental health[53]. Conversely, the male profile indicates an externalizing pathway, with physical fighting’s persistent high importance (SHAP +0.034 to +0.036) serving as a behavioral marker for conduct problems and interpersonal violence common in males[54]. The critical contribution of our study lies in dynamically modeling risk architectures across adolescence, providing granular evidence of developmental trajectories. For boys, problematic social media use importance declines while academic pressure rises with age, suggesting developmental shifts from early-adolescent behavioral dysregulation toward later-adolescent societal and academic pressures. Female profile stability may reflect persistent socio-evaluative pressures intensifying at puberty[41-43, 52]. These distinct pathways argue against “one-size-fits-all” approaches, advocating developmentally timed, sex-specific strategies: for girls, managing academic anxiety and fostering digital citizenship; for boys, emphasizing emotional regulation and pro-social conflict resolution.

### Implications

This study provides a multi-faceted, data-driven framework with actionable implications for key stakeholders in adolescent well-being. For Policymakers, the research offers a methodological blueprint for data-driven mental health policies. Machine learning models achieving high accuracy (ROC 0.76-0.79) using existing survey data demonstrate that instruments like HBSC can transform from descriptive tools into predictive engines for population-level surveillance. This capability enables effective resource targeting for large-scale initiatives, such as the European Commission’s EUR 1.23 billion mental health strategy. SHAP analysis reveals a quantitative hierarchy identifying a “globalized signature” of adolescent stress, with academic pressure and problematic social media use as dominant predictors across 46 countries[48], providing evidence-based mandate for cross-sectoral investment in educational reforms and digital wellness programs. Distinct sex-specific risk profiles necessitate a “glocal” policy approach addressing global stressors while tailoring interventions to sex and developmental stage. For Clinicians and Public Health Practitioners, the model serves as validated clinical decision support for early identification, flagging at-risk adolescents who may not meet diagnostic thresholds or present for care. SHAP analysis provides a dynamic “map” of distress combating diagnostic overshadowing, prompting clinicians to probe underlying psychosocial drivers when patients present with somatic complaints. This enables nuanced assessment frameworks attuned to age- and sex-specific risk factors. The methodology exemplifies responsible AI application through non-interactive analytical engines, explainable AI transparency, and human-in-the-loop interpretation, aligning with ethical guidelines while augmenting rather than replacing professional care. For Teachers and School Administrators, findings offer concrete strategies for evidence-based support systems. The predictive model suits universal screening within Multi-Tiered Support Systems, enabling proactive identification and precise resource allocation. Academic pressure’s emergence as the dominant predictor challenges schools to re-evaluate learning environments as primary health determinants, reframing issues from individual deficits to systemic problems requiring “whole-school” policy changes. The study reinforces fostering protective factors like teacher and peer support while implementing comprehensive digital citizenship curricula. For Parents and Caregivers, research provides empowering guidance emphasizing family support and open communication as powerful buffers against external stressors[49]. The study offers nuanced technology navigation frameworks, identifying problematic social media use—characterized by addiction-like symptoms rather than screen time—suggesting collaborative, relational approaches focused on digital literacy and emotional processing. Awareness of sex-specific warning signs (internalizing symptoms in girls versus externalizing behaviors in boys) enables better recognition of emerging distress and timely support-seeking.

### Strength and limitations

This investigation demonstrates several key strengths establishing it as a significant contribution to adolescent mental health surveillance. The principal strength lies in unprecedented scale and geographic scope, leveraging HBSC survey data from 225,000 adolescents across 46 countries. This cross-national dataset provides statistical power and generalizability rare in mental health prediction research, which has typically been confined to single populations or clinical settings. The robust discriminatory performance (ROC 0.76-0.79) demonstrates that standardized, cost-effective survey instruments can achieve predictive accuracy approaching resource-intensive clinical data sources like electronic health records (ROC 0.78-0.81)[15]. This provides a methodological blueprint for transforming descriptive public health tools into predictive engines for global surveillance. Methodologically, systematic comparison of eight machine learning algorithms enhances findings’ robustness, while Shapley Additive Explanations (SHAP) ensures model interpretability, advancing beyond “black-box” prediction toward trustworthy, explainable AI. This transparency is crucial for clinical adoption and ethical deployment[55]. Stratification by sex and grade represents a core strength, uncovering novel insights into dynamic, sex-differentiated risk architectures. By providing large-scale quantitative validation for established internalizing versus externalizing pathways in psychopathology, the study offers compelling, data-driven rationale for designing developmentally-timed and sex-specific public health strategies.

Despite these strengths, the study is subject to several limitations that frame a clear agenda for future research. The primary limitation is the cross-sectional nature of the HBSC data, which, despite its scale, precludes the establishment of causal relationships. While the models identify powerful predictive associations, they cannot determine temporal precedence; for instance, it remains unclear whether academic pressure causes psychosomatic complaints or if adolescents prone to such complaints are more sensitive to academic pressure. Future research must therefore apply these models to longitudinal cohort data to disentangle these complex, bidirectional relationships and validate the developmental trajectories suggested by the current findings[7, 38]. A second limitation is the exclusive reliance on adolescent self-report, which introduces susceptibility to measurement biases such as social desirability and recall error[56, 57]. Critically, this single-informant approach overlooks the well-documented phenomenon of parent-adolescent discrepancy in reporting internalizing symptoms, where adolescents often report higher symptom levels than their parents[58]. Future models should incorporate multi-informant data and could benefit from explicitly modeling the informant discrepancy itself as a predictor of psychopathology. Furthermore, while the 46-country sample is expansive, it is predominantly composed of countries that are Western, Educated, Industrialized, Rich, and Democratic (WEIRD). As research by Henrich et al. (2010) suggests, findings from WEIRD populations are often psychological outliers and may not be representative of the human species[59]. Consequently, the identified “globalized signature” of adolescent stress may not generalize to non-WEIRD cultural contexts where constructs like academic pressure and family support hold different meanings[60]. It is therefore imperative to validate and retrain these models on data from diverse, non-WEIRD populations to develop culturally sensitive prediction tools. Finally, while the use of SHAP is a strength, its interpretation requires caution, as SHAP values explain the model’s output, not necessarily the real-world causal mechanism, and can perpetuate any biases inherent in the underlying algorithm.

## Conclusion

This investigation provides robust, multi-national validation of a machine learning framework for early identification of adolescents at high mental health risk. By demonstrating that scalable survey data can achieve predictive performance (ROC 0.76-0.79) comparable to clinical data sources, this study establishes a viable paradigm for population-level mental health surveillance. The research advances beyond prediction through explainable AI, revealing an evidence-based hierarchy where academic pressure and problematic social media use emerge as dominant risks, while family support serves as the most potent protective factor. The findings challenge “one-size-fits-all” approaches by elucidating distinct, sex-differentiated risk architectures. Physical fighting’s unique salience for boys versus relational and academic pressures for girls provides compelling rationale for precisely tailored interventions: managing academic anxiety and fostering digital citizenship for girls, while emphasizing emotional regulation and prosocial conflict resolution for boys. Future research should integrate longitudinal data to establish causality, incorporate multi-modal data streams including digital biomarkers, and ensure global equity through cross-cultural validation in non-WEIRD populations. This integrated agenda can transition the field from risk prediction to mechanistic understanding, ultimately realizing precision public health for adolescent mental well-being.

## Contributors

SC contributed to the concept and study design. SC conducted the analysis. SC, CL, YC, SD, YY, YL, JL, GL, QAL, and MN made critical revisions to the manuscript for important intellectual content. All authors edited and approved the final manuscript.

## Funding

SC’s research was supported by the PENDA, funded by the UK Foreign, Commonwealth and Development Office.

## Role of the funding source

The funder of the study had no role in study design, data collection, data analysis, data interpretation, or writing of the article. For the purpose of open access, the authors have applied a Creative Commons Attribution (CC BY) licence to any Author Accepted Manuscript version arising from this submission.

## Conflict of Interest

SC and other authors declare no conflict of interest with this work.

## Ethics Statement

The use of secondary de-identified data makes this study exempt from institutional review board review. Each original survey of HBSC received ethics approval from an appropriate regulatory body in each participating country, and informed consent was obtained from participants and their parents.

## Data Availability Statement

The data are publicly available and can be accessed here (https://hbsc.org/data/).

## Notes

### Competing Interest Statement

The authors have declared no competing interest.

### Funding Statement

SC's research was supported by the PENDA, funded by the UK Foreign, Commonwealth and Development Office. The funder of the study had no role in study design, data collection, data analysis, data interpretation, or writing of the article.

### Author Declarations

The study used ONLY openly available human data that were originally located at https://hbsc.org/data/.

