## Supplementary material for "Identifying High-Risk Adolescents for Mental Health Difficulties: A Machine Learning Analysis of the Health Behaviour in School-aged Children Study Across 46 Countries": Tables and figures

**Table 1. Basic description**. Data were presented as mean (standard deviation [SD]) for continuous variables or number (percentage) for categorical variables. Between-group balance for multiple psychosomatic health complaints (MPHC) was assessed using standardized mean differences (SMD), where SMD ≥ 0.1 indicated meaningful imbalance between groups.

| **Variables** | **MPHC (= No) (N = 190587)** | **MPHC (= Yes) (N = 34834)** | **All (N = 225421)** | **SMD** |
| --- | --- | --- | --- | --- |
| **Age (in years)** | 13.48 (1.63) | 13.75 (1.63) | 13.52 (1.63) | -0.17 |
| **Sex** |  |  |  |  |
| Boy | 97675 (51.2%) | 12370 (35.5%) | 110045 (48.8%) | -0.32 |
| Girl | 92912 (48.8%) | 22464 (64.5%) | 115376 (51.2%) |  |
| **Grade** |  |  |  |  |
| Grade 5 | 67647 (35.5%) | 10236 (29.4%) | 77883 (34.6%) | 0.15 |
| Grade 7 | 64097 (33.6%) | 11826 (33.9%) | 75923 (33.7%) |  |
| Grade 9 | 58843 (30.9%) | 12772 (36.7%) | 71615 (31.8%) |  |
| **Family affluence** | 7.99 (2.73) | 7.51 (2.80) | 7.92 (2.74) | 0.17 |
| **Living with mother (=yes)** | 178874 (93.9%) | 31688 (91.0%) | 210562 (93.4%) | 0.11 |
| **Living with father (=yes)** | 142253 (74.6%) | 22509 (64.6%) | 164762 (73.1%) | 0.22 |
| **Living in child home (=yes)** | 1603 (0.8%) | 551 (1.6%) | 2154 (1.0%) | -0.07 |
| **Body perception** |  |  |  |  |
| Much too thin | 5711 (3.0%) | 2022 (5.8%) | 7733 (3.4%) | 0.42 |
| A bit too thin | 26840 (14.1%) | 4632 (13.3%) | 31472 (14.0%) |  |
| About right | 111413 (58.5%) | 14695 (42.2%) | 126108 (55.9%) |  |
| A bit too fat | 41683 (21.9%) | 10334 (29.7%) | 52017 (23.1%) |  |
| Much too fat | 4940 (2.6%) | 3151 (9.0%) | 8091 (3.6%) |  |
| **Body mass index z-scores** |  |  |  |  |
| Thinness | 12541 (6.6%) | 2483 (7.1%) | 15024 (6.7%) | 0.07 |
| Underweight | 26624 (14.0%) | 4571 (13.1%) | 31195 (13.8%) |  |
| Normal weight | 109514 (57.5%) | 19356 (55.6%) | 128870 (57.2%) |  |
| Overweight | 30935 (16.2%) | 5989 (17.2%) | 36924 (16.4%) |  |
| Obese | 10973 (5.8%) | 2435 (7.0%) | 13408 (5.9%) |  |
| **Days smoked in the last 30 days** |  |  |  |  |
| Never | 179298 (94.1%) | 29994 (86.1%) | 209292 (92.8%) | 0.27 |
| 1-2 days | 4082 (2.1%) | 1552 (4.5%) | 5634 (2.5%) |  |
| 3-5 days | 1595 (0.8%) | 604 (1.7%) | 2199 (1.0%) |  |
| 6-9 days | 1104 (0.6%) | 401 (1.2%) | 1505 (0.7%) |  |
| 10-19 days | 1059 (0.6%) | 462 (1.3%) | 1521 (0.7%) |  |
| 20-29 days | 933 (0.5%) | 375 (1.1%) | 1308 (0.6%) |  |
| 30-31 days | 2516 (1.3%) | 1446 (4.2%) | 3962 (1.8%) |  |
| **Alcohol use in the last 30 days** |  |  |  |  |
| Never | 156737 (82.2%) | 24859 (71.4%) | 181596 (80.6%) | 0.28 |
| 1-2 days | 21514 (11.3%) | 5582 (16.0%) | 27096 (12.0%) |  |
| 3-5 days | 6516 (3.4%) | 1963 (5.6%) | 8479 (3.8%) |  |
| 6-9 days | 2832 (1.5%) | 1025 (2.9%) | 3857 (1.7%) |  |
| 10-19 days | 1409 (0.7%) | 523 (1.5%) | 1932 (0.9%) |  |
| 20-29 days | 399 (0.2%) | 227 (0.7%) | 626 (0.3%) |  |
| 30-31 days | 1180 (0.6%) | 655 (1.9%) | 1835 (0.8%) |  |
| **Cannabis in the last 30 days** |  |  |  |  |
| Never | 183282 (96.2%) | 32393 (93.0%) | 215675 (95.7%) | 0.14 |
| 1-2 days | 3342 (1.8%) | 1092 (3.1%) | 4434 (2.0%) |  |
| 3-5 days | 1158 (0.6%) | 388 (1.1%) | 1546 (0.7%) |  |
| 6-9 days | 793 (0.4%) | 235 (0.7%) | 1028 (0.5%) |  |
| 10-19 days | 699 (0.4%) | 211 (0.6%) | 910 (0.4%) |  |
| 20-29 days | 355 (0.2%) | 103 (0.3%) | 458 (0.2%) |  |
| 30-31 days | 958 (0.5%) | 412 (1.2%) | 1370 (0.6%) |  |
| **Physical activity in the past 7 days (days)** | 4.14 (2.04) | 3.65 (2.26) | 4.06 (2.08) | 0.23 |
| **Physical activity out of school in the past 7 days (days)** | 2.99 (1.59) | 3.33 (1.88) | 3.04 (1.64) | -0.20 |
| **Support from friends** | 21.34 (6.88) | 19.46 (7.66) | 21.05 (7.04) | 0.26 |
| **Support from family** | 23.16 (6.73) | 20.01 (7.64) | 22.67 (6.97) | 0.44 |
| **Support from students** | 11.69 (2.43) | 10.51 (2.98) | 11.51 (2.56) | 0.43 |
| **Support from teachers** | 11.63 (2.64) | 10.45 (3.21) | 11.45 (2.77) | 0.40 |
| **Talk to father** |  |  |  |  |
| Very easy | 64510 (33.8%) | 8344 (24.0%) | 72854 (32.3%) | 0.48 |
| Easy | 68984 (36.2%) | 8591 (24.7%) | 77575 (34.4%) |  |
| Difficult | 32356 (17.0%) | 7840 (22.5%) | 40196 (17.8%) |  |
| Very difficult | 12838 (6.7%) | 6317 (18.1%) | 19155 (8.5%) |  |
| Don`t have or see | 11899 (6.2%) | 3742 (10.7%) | 15641 (6.9%) |  |
| **Talk to stepfather** |  |  |  |  |
| Very easy | 9001 (4.7%) | 1624 (4.7%) | 10625 (4.7%) | 0.21 |
| Easy | 13616 (7.1%) | 2203 (6.3%) | 15819 (7.0%) |  |
| Difficult | 11079 (5.8%) | 2392 (6.9%) | 13471 (6.0%) |  |
| Very difficult | 8167 (4.3%) | 3258 (9.4%) | 11425 (5.1%) |  |
| Don`t have or see | 148724 (78.0%) | 25357 (72.8%) | 174081 (77.2%) |  |
| **Talk to mother** |  |  |  |  |
| Very easy | 99432 (52.2%) | 13683 (39.3%) | 113115 (50.2%) | 0.44 |
| Easy | 64172 (33.7%) | 10246 (29.4%) | 74418 (33.0%) |  |
| Difficult | 17940 (9.4%) | 6030 (17.3%) | 23970 (10.6%) |  |
| Very difficult | 5668 (3.0%) | 3671 (10.5%) | 9339 (4.1%) |  |
| Don`t have or see | 3375 (1.8%) | 1204 (3.5%) | 4579 (2.0%) |  |
| **Talk to stepmother** |  |  |  |  |
| Very easy | 9077 (4.8%) | 1664 (4.8%) | 10741 (4.8%) | 0.18 |
| Easy | 11175 (5.9%) | 1822 (5.2%) | 12997 (5.8%) |  |
| Difficult | 9700 (5.1%) | 2036 (5.8%) | 11736 (5.2%) |  |
| Very difficult | 7788 (4.1%) | 2855 (8.2%) | 10643 (4.7%) |  |
| Don`t have or see | 152847 (80.2%) | 26457 (76.0%) | 179304 (79.5%) |  |
| **Digital communication preference** | 6.43 (3.66) | 7.24 (4.14) | 6.56 (3.75) | -0.21 |
| **Intense social media use** | 13.39 (4.57) | 13.94 (4.94) | 13.48 (4.64) | -0.12 |
| **Problematic social media use** | 1.61 (2.00) | 2.67 (2.45) | 1.77 (2.11) | -0.47 |
| **Been bullied in the past months** |  |  |  |  |
| Haven't | 144269 (75.7%) | 20449 (58.7%) | 164718 (73.1%) | 0.42 |
| Once or twice | 30034 (15.8%) | 7071 (20.3%) | 37105 (16.5%) |  |
| 2-3 times per month | 7037 (3.7%) | 2477 (7.1%) | 9514 (4.2%) |  |
| Once/week | 4353 (2.3%) | 1645 (4.7%) | 5998 (2.7%) |  |
| Several times/week | 4894 (2.6%) | 3192 (9.2%) | 8086 (3.6%) |  |
| **Been cyberbullied in the past months** |  |  |  |  |
| Haven't | 169713 (89.0%) | 26253 (75.4%) | 195966 (86.9%) | 0.37 |
| Once or twice | 14862 (7.8%) | 5162 (14.8%) | 20024 (8.9%) |  |
| 2-3 times per month | 3042 (1.6%) | 1488 (4.3%) | 4530 (2.0%) |  |
| Once/week | 1385 (0.7%) | 788 (2.3%) | 2173 (1.0%) |  |
| Several times/week | 1585 (0.8%) | 1143 (3.3%) | 2728 (1.2%) |  |
| **Physical fighting in the past 12 months** |  |  |  |  |
| None | 126866 (66.6%) | 19286 (55.4%) | 146152 (64.8%) | 0.29 |
| 1 time | 31030 (16.3%) | 6068 (17.4%) | 37098 (16.5%) |  |
| 2 times | 14611 (7.7%) | 3227 (9.3%) | 17838 (7.9%) |  |
| 3 times | 6924 (3.6%) | 1907 (5.5%) | 8831 (3.9%) |  |
| 4 times or more | 11156 (5.9%) | 4346 (12.5%) | 15502 (6.9%) |  |
| **Injured in the past 12 months** |  |  |  |  |
| None | 110097 (57.8%) | 16862 (48.4%) | 126959 (56.3%) | 0.29 |
| 1 time | 43060 (22.6%) | 7371 (21.2%) | 50431 (22.4%) |  |
| 2 times | 19956 (10.5%) | 4377 (12.6%) | 24333 (10.8%) |  |
| 3 times | 8646 (4.5%) | 2413 (6.9%) | 11059 (4.9%) |  |
| 4 times or more | 8828 (4.6%) | 3811 (10.9%) | 12639 (5.6%) |  |
| **Liking school** |  |  |  |  |
| Like a lot | 55629 (29.2%) | 6929 (19.9%) | 62558 (27.8%) | 0.47 |
| Like a bit | 89780 (47.1%) | 12782 (36.7%) | 102562 (45.5%) |  |
| Not very much | 32511 (17.1%) | 8405 (24.1%) | 40916 (18.2%) |  |
| Not at all | 12667 (6.6%) | 6718 (19.3%) | 19385 (8.6%) |  |
| **Perceived academic pressure** |  |  |  |  |
| Not at all | 42550 (22.3%) | 5045 (14.5%) | 47595 (21.1%) | 0.56 |
| A little | 85050 (44.6%) | 10310 (29.6%) | 95360 (42.3%) |  |
| Some | 43450 (22.8%) | 9194 (26.4%) | 52644 (23.4%) |  |
| A lot | 19537 (10.3%) | 10285 (29.5%) | 29822 (13.2%) |  |

**Figure 1**. **Receiver operating characteristic (ROC) curves for machine learning models predicting multiple psychosomatic health complaints in children.** ROC curves demonstrate the predictive performance of optimal models selected from eight machine learning approaches. The x-axis represents the false positive rate (1 - specificity) and the y-axis represents the true positive rate (sensitivity). Each panel displays the best-performing model within its respective modeling framework, as determined by the highest area under the ROC curve value. The legend presents ROC values with their corresponding 95% confidence intervals (CI).

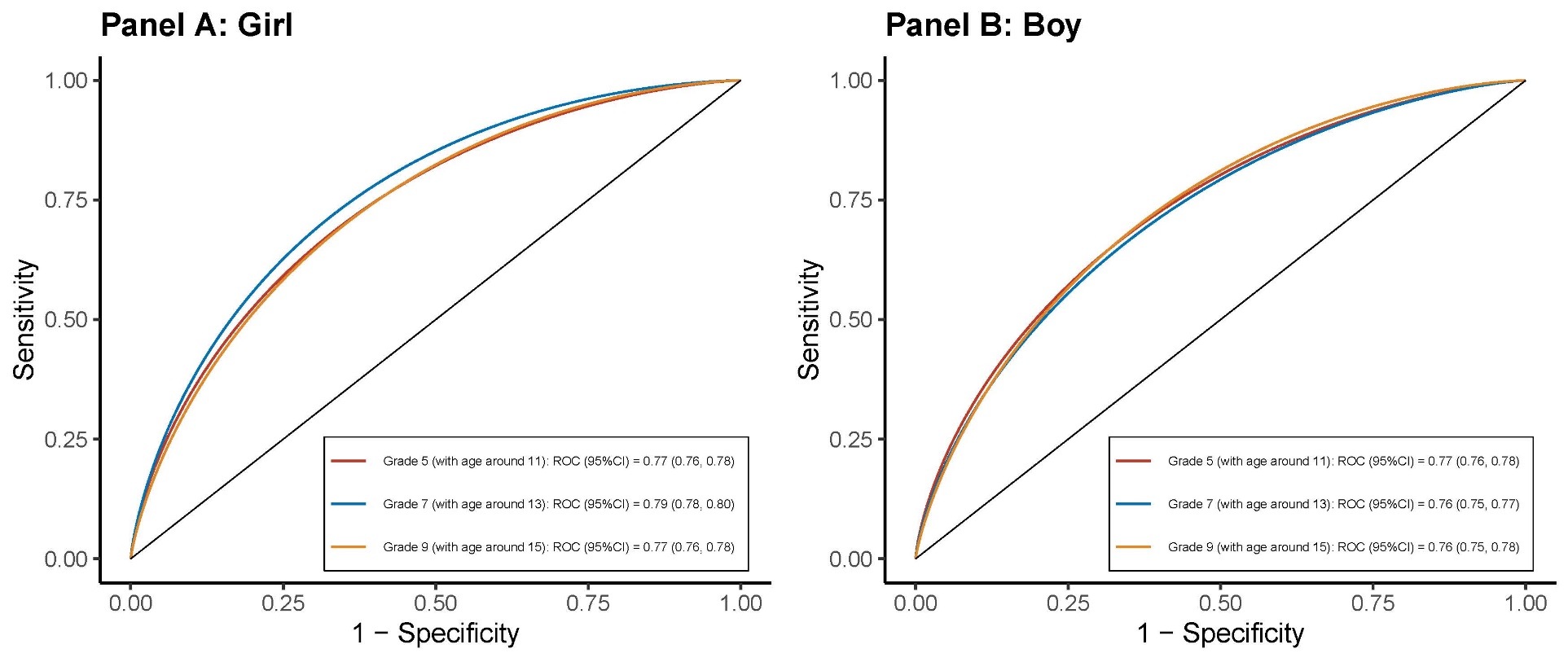

**Figure 2**. **SHAP analysis of optimal machine learning model predictions stratified by sex and grade level.** This figure presents SHapley Additive exPlanations (SHAP) analysis results for the best-performing machine learning model. Each horizontal row represents an individual feature, while the x-axis displays SHAP values quantifying each feature's impact on model predictions. Data points are color-coded by feature magnitude, with red indicating higher values and blue indicating lower values, visualizing both the direction and strength of feature influence on model outcomes.

| 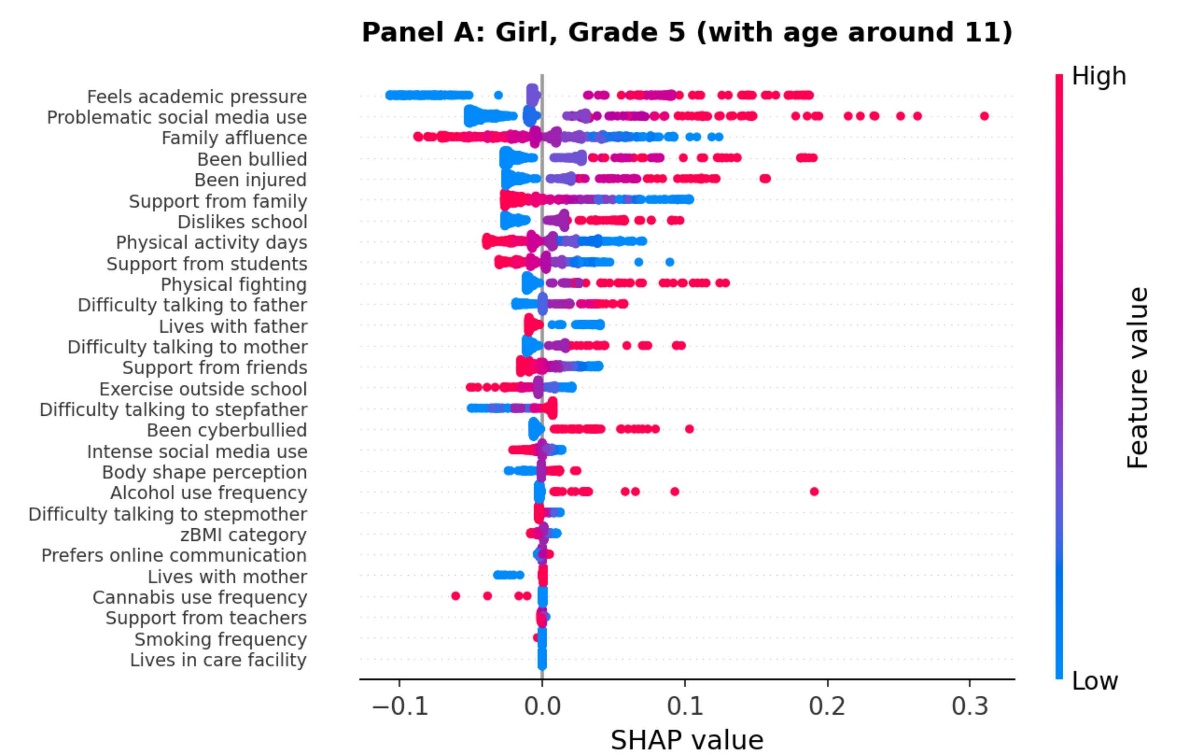 | 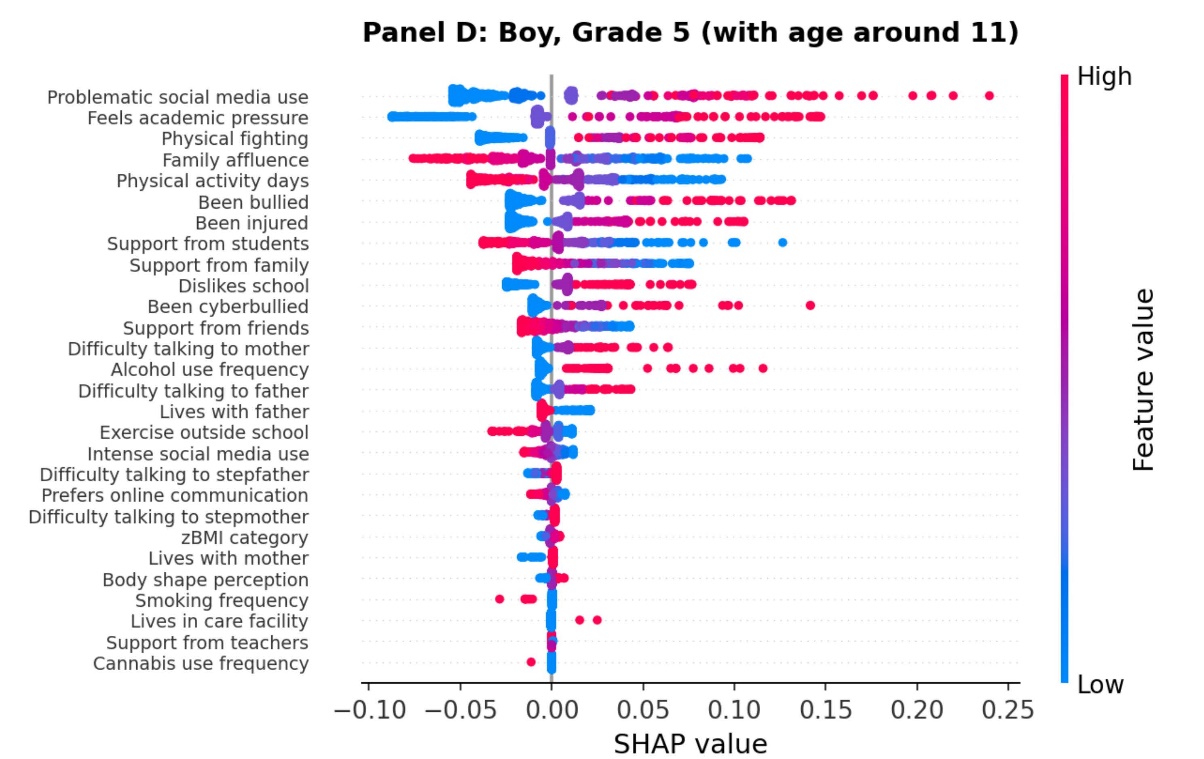 |
| --- | --- |
| 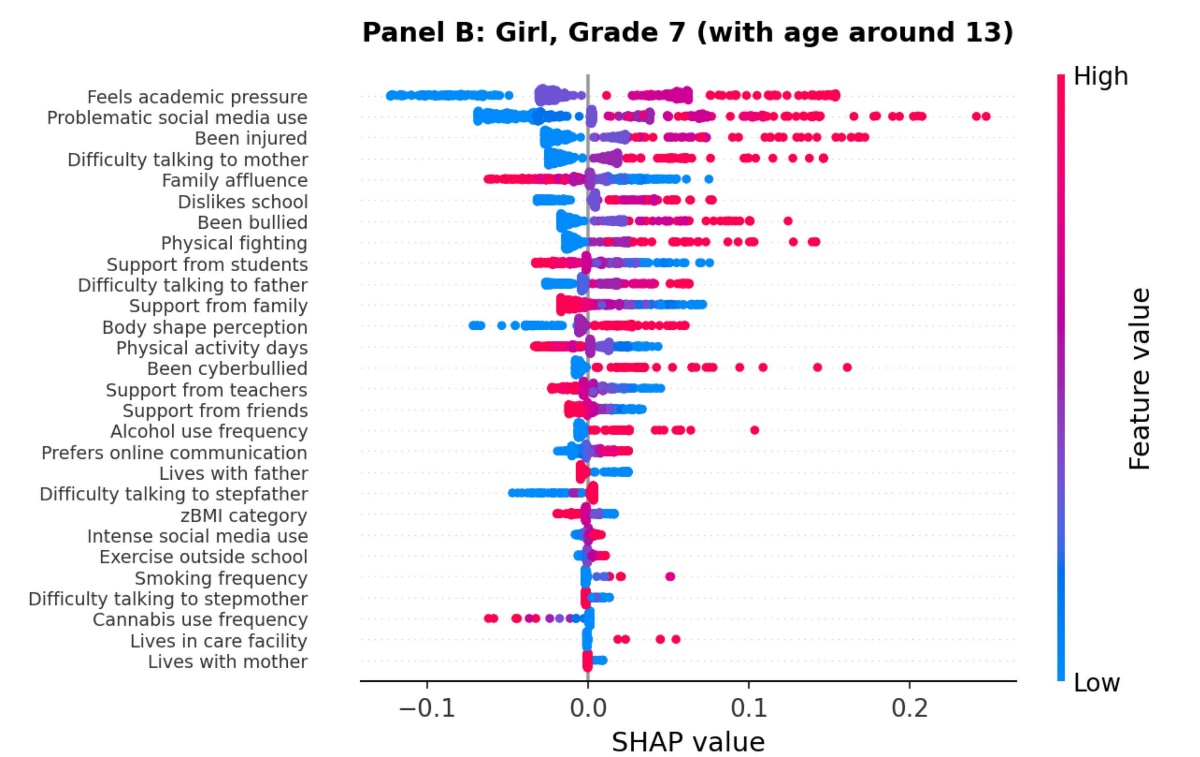 | 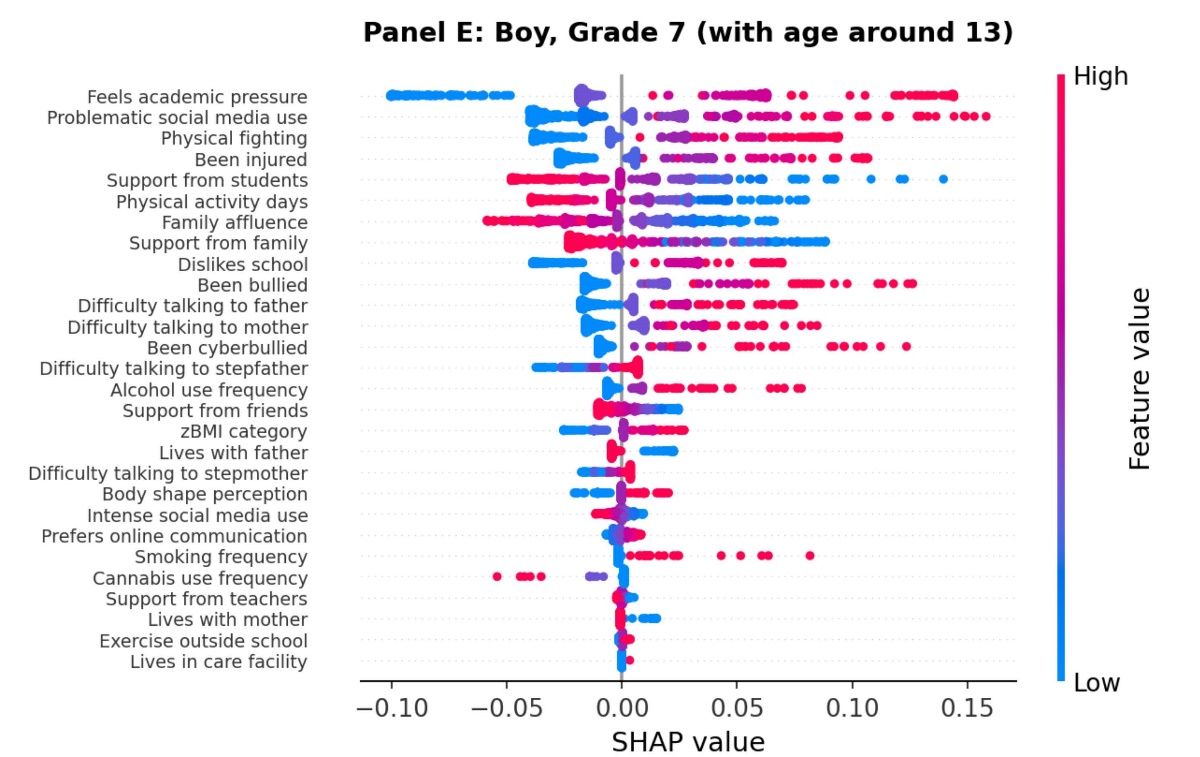 |
| 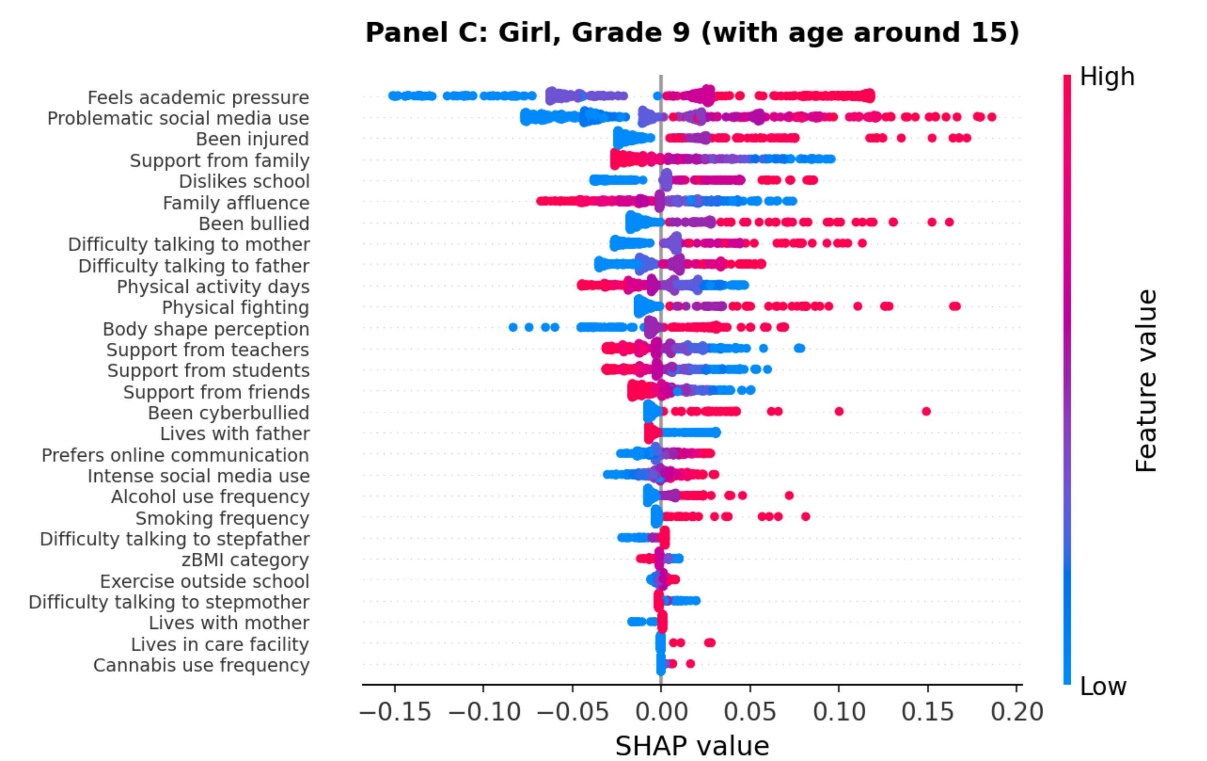 | 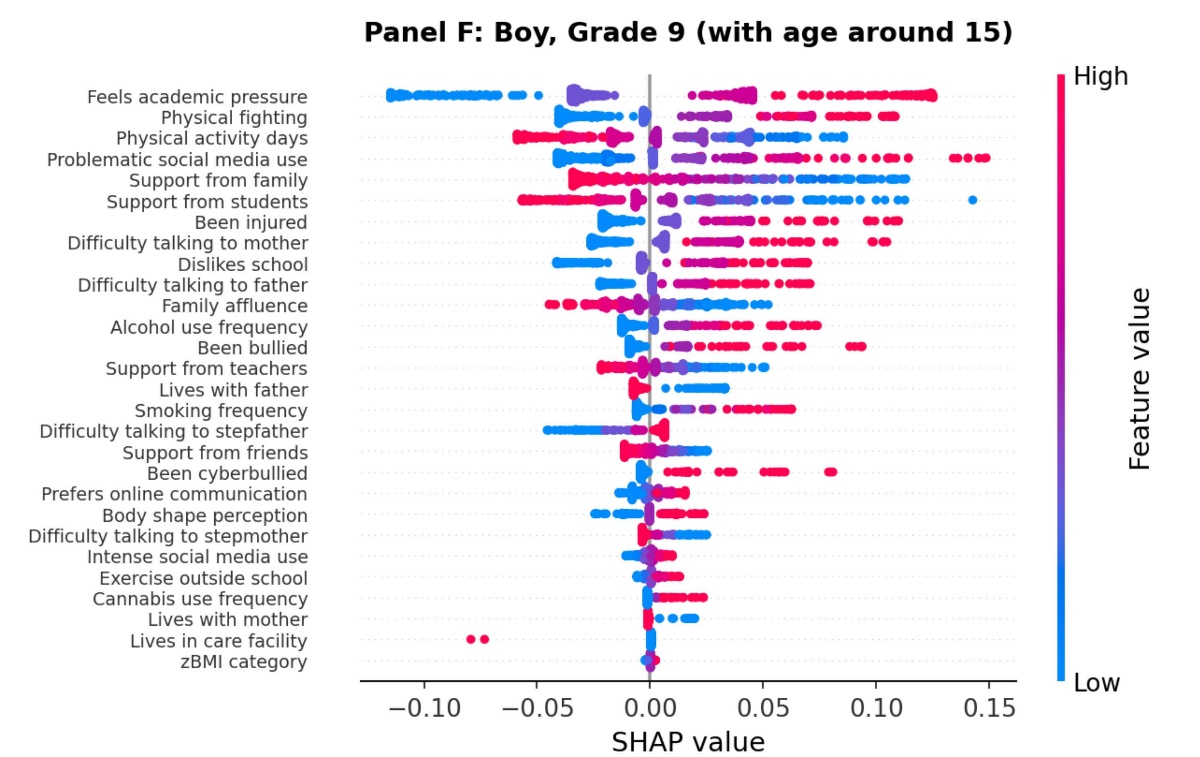 |

**Figure 3.** **Feature importance rankings for optimal machine learning models stratified by sex and grade level**. This figure displays the relative importance of predictive features as determined by the best-performing machine learning model within each demographic subgroup. Feature importance scores quantify each variable's contribution to model predictive accuracy, with the top 15 most influential predictors shown for each sex-grade combination. Red bars indicate features that increase the risk of multiple psychosomatic health complaints, while blue bars represent protective factors that decrease risk. The connecting curves between panels illustrate how feature importance changes across grade levels, with red lines indicating increasing importance and blue lines indicating stable or decreasing importance over time.
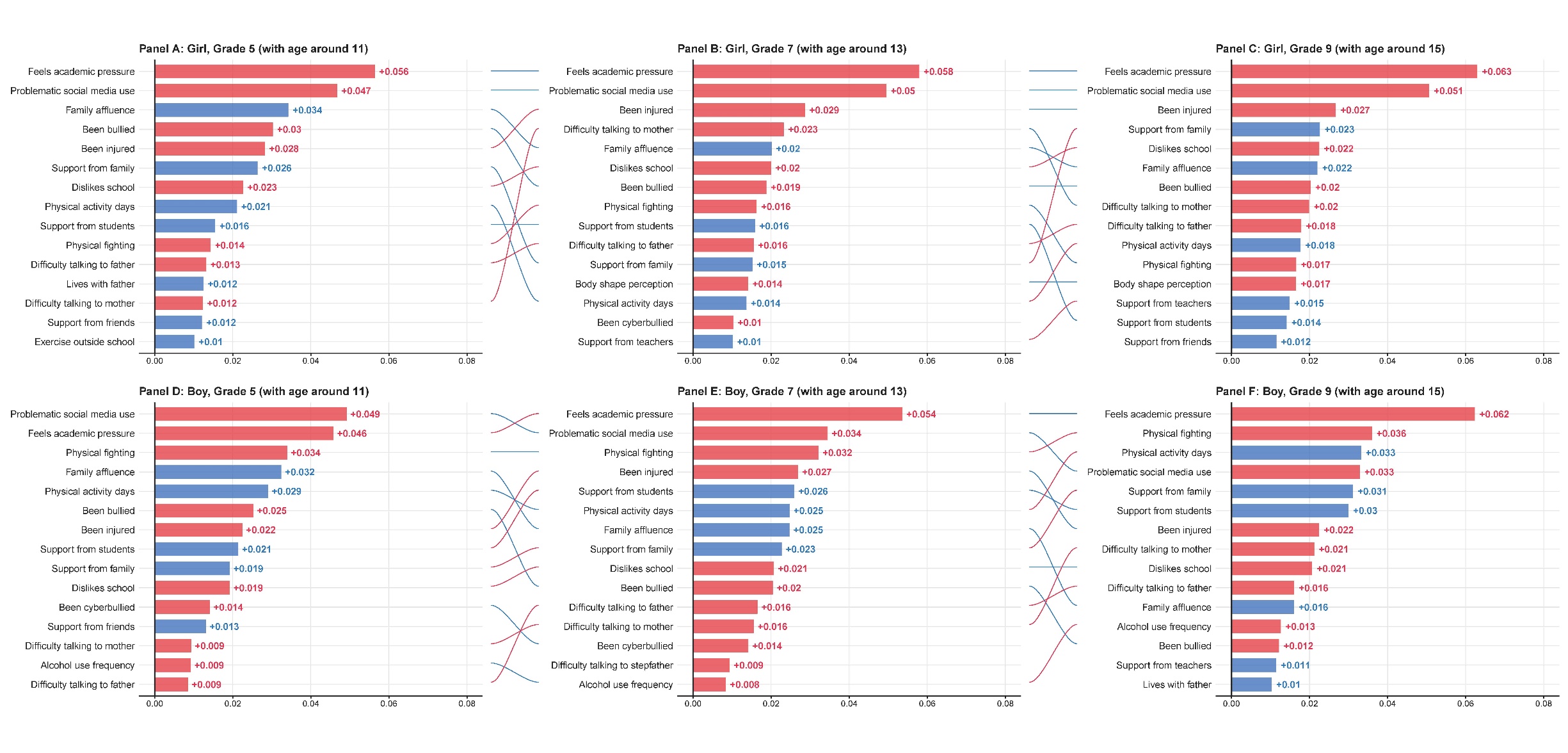
